# Cannabis Use Patterns in First Episode Psychosis and Schizophrenia: A Scoping Review and Case Series

**DOI:** 10.1101/2025.04.28.25325846

**Authors:** Jeff W. Jin, Nolan J. Neu, Isaac B. Satz, Janelle Annor, Mohamed ElSayed, Mary F. Brunette

## Abstract

**Background:** Cannabis use is associated with psychosis development and symptom relapse in persons with schizophrenia spectrum disorders (SCZ). As more U.S. states legalize cannabis and products increase in potency, it is crucial to better understand recent cannabis use patterns in SCZ.

**Methods:** We conducted a scoping review of research on cannabis use patterns in SCZ after January 2016 and present a case series of cannabis use in six inpatients with psychosis from 2023–2024.

**Results:** Scoping review: Of 672 references, nine studies (775 participants) were included; none were designed to characterize cannabis quantity, frequency, or type of use over time. Cannabis measurement methodology varied widely and most studies did not follow recommendations for measuring cannabis use. Frequency and quantity of use at study baseline were reported by most studies and these ranged widely. At least a minority of participants with SCZ in each study used cannabis very frequently; quantity of used ranged widely from 0.6±0.6 to 3.4±2.2 joints/day. One small study detailed cannabis product type among users for THC (93% flower, 80% edibles, 60% concentrates) and CBD (40% flower, 20% edibles, 20% concentrates, 13% oils).

**Case Series:** Participants were inpatients (32.0±14.4 years; 83.3% diagnosed with SCZ) who used cannabis 2.7±2.1 days/week. All used cannabis leaf (3.1±2.3 joints/day); half (all heavy users) also used concentrates (33.3%) or edibles (16.7%).

**Conclusion:** Only nine recent studies measured cannabis use patterns in SCZ; methodologies varied. As cannabis legalization expands and product potency increases, further research should characterize cannabis use and its consequences in SCZ.

## 1. Introduction

Schizophrenia Spectrum Disorders (SCZ – including delusional, brief psychotic, schizophreniform, schizoaffective, schizophrenia, and unspecified psychotic disorders) are prevalent in 0.5–1% of the population (Rössler et al., 2005) and carry a high risk of co-morbid substance use disorders (SUD) (Hunt et al., 2018). Cannabis use disorder (CUD) is the second most common co-occurring SUD in SCZ, with a recent world prevalence of 26.2% and North American prevalence of 23.5% (Hunt et al., 2018). Prevalences reported over the past 25 years have ranged from 13% to 45% (Koskinen et al., 2009). In contrast, the estimated recent prevalence of CUD in the U.S. general population is 1–6% (Hasin et al., 2016; Hasin et al., 2019).

Cannabis use in SCZ has been associated with symptom exacerbation, medication non-adherence, and hospitalizations over time (Foglia et al., 2017; Hasan et al., 2020; Schoeler et al., 2017). In observational research and a small number of single dose, placebo-controlled trials, the main psychoactive component of cannabis, tetrahydrocannabinol (THC), was associated with worse cognitive, positive, and negative symptoms in SCZ in a dose-dependent manner (Ahmed et al., 2021; Brunette et al., 2024; Murray et al., 2017; Stark et al., 2021). In SCZ patients with cannabis use (with or without CUD), cannabis reduction or abstinence was associated with reduced symptoms or relapses of psychosis over time (Levi et al., 2023; Marino et al., 2020; Schoeler et al., 2016; Schoeler et al., 2017; Sheitman et al., 2024), long-term improvements in negative psychotic symptoms (González-Pinto et al., 2011), and increases in cognitive function – particularly memory and verbal learning (Rabin et al., 2017). In healthy individuals, high levels of cannabis use may increase the risk for psychosis and for the development of SCZ (D’Souza et al., 2022; Groening et al., 2024; Hasan et al., 2020). In fact, a notable recent study of emergency and hospital admission data in Ontario, Canada showed that the population–attributable fraction (PARF) of CUD associated with SCZ more than doubled since cannabis was legalized. In contrast, cannabidiol (CBD), a non-psychoactive component of cannabis, does not appear to be associated with psychosis (Ahmed et al, 2021) and may attenuate the effects of THC on psychosis and some cognitive symptoms (Bartoli et al., 2021; Lawn et al., 2016). Thus, the current evidence suggests that people with psychotic disorders are a vulnerable population that may be harmed by using substances or products containing THC. However, the harmful amount of use (dose and/or frequency) is not clearly established.

In 2017, there were an estimated 188 million adult cannabis users (3.8%) across the globe (UNODC, 2019). In the U.S., the prevalence of cannabis use has increased over the past 20 years (Chiu et al., 2021) across most age, race, and ethnic groups (Patrick et al., 2024), with 19.4% of adults reporting past–year use in 2022–2023 (SAMHSA, 2023). In 2022, cannabis was even used more than alcohol in the U.S. (Caulkins, 2024). During this time, laws have changed to either decriminalize or legalize the use of “medical” or recreational cannabis in more than 24 states (NCSL, 2024). Concurrently, the sale and use of cannabis products have risen steadily, cannabis product concentrations (street and legally obtained) have increased, and cannabis prices have decreased (Hasin et al., 2019; Hinckley et al., 2024). Legalization has been associated with increased prevalence of co–use smoking of cannabis leaf products and use of other products with higher concentrations (Borodovsky et al., 2023; Hasin et al., 2019; Manthey et al., 2021). Furthermore, most legally sold and street–obtained products now have THC concentrations greater than 15% (Chiu et al., 2021; ElSohly et al., 2021; Smart et al., 2017). Although these higher concentration products have greater potential to contribute to the development of CUD (Arterberry et al., 2019) or induce psychosis (D’Souza et al., 2022) compared to the lower potency products that were predominant in the past, public perceptions of risk or harm have decreased (Florimbio et al., 2023; Lemos et al., 2023).

Given increasing legalization, perceptions of reduced harm, and the known impacts of cannabis on SCZ, it is imperative to better understand patterns of cannabis use in this vulnerable population in the new landscape of easier availability and higher potency. A 2018 online survey aimed to capture this by examining cannabis use patterns in a non-clinical Canadian and U.S. sample selected from non-probability commercial panels recruited via email. Their findings showed that persons with self-reported psychosis were more likely to report frequent, heavy cannabis use and used potentially more potent oral and topical products compared to those without self-reported psychosis (Rup et al., 2021). Specifically, 20.3% of people with self-reported psychosis reported past year daily use, 13.6% reported weekly to monthly use, 9.8% reported less than monthly cannabis use, and 55.3% did not use cannabis. Smoking dried cannabis herb (76–86%) and using edibles (36–46%) were common modalities. Other products were used less frequently – vape oil (24–35%), oral oils (17–27%), beverages (8– 11%), and tinctures (5–10%). This study provided insight into cannabis use patterns reported by survey respondents in the general population, but its commercial survey panel sample is likely different from people in services for the treatment of SCZ.

Therefore, our study used a scoping review methodology to examine currently available research on quantity, frequency, and type of cannabis use in people clinically diagnosed with SCZ in the context of state cannabis legalization in the U.S. We also present a descriptive case series of past–month cannabis use patterns in recently admitted psychiatric inpatients with psychosis during 2023–2024.

## 2. Methods

### 2.1 Literature Search Strategy and Article Screening

We searched the PubMed database to identify relevant studies published between January 2016 and February 2024 as legalization was underway in eight states by 2016 and expanded dramatically thereafter. The search strategy focused on subject headings and related terms using variations of “Cannabis” and “Schizophrenia” with various additional descriptors of cannabis use characteristics and outcomes such as “potency” and “legalization.” Further detailed information on the search strategy is in the online supplement. Two reviewers (JJ, NN) independently screened the search results by title, abstract, and eligibility using Rayyan QCRI software (Ouzzani et al., 2016). Disagreements were resolved through discussion with a third investigator (MB) and consensus agreement. Publication data extraction was performed independently by two reviewers (JJ, NN) and all data was checked by a third team member (MB).

### 2.2 Literature Review Inclusion Criteria

The major inclusion criteria were: [1] publication in English language, [2] inclusion of individuals with Diagnostic and Statistical Manual of Mental Disorders, Fourth Edition (DSM-IV), or revised diagnosis of SCZ (schizophrenia, schizoaffective disorder, schizophreniform disorder) including chronic SCZ, first episode psychosis [FEP], or recent-onset psychosis [ROP]), [3] recruitment of subjects resulting in sample with at least half of data collected after 2016, and [4] systematic assessment of cannabis use, frequency, or other characteristics. Study designs with retrospective and prospective assessments were allowed. We included studies that recruited individuals with other psychiatric diagnostic groups (in addition to SCZ) if SCZ reflected a majority of the sample or if cannabis characteristics were reported separately for SCZ and other diagnosis groups.

Exclusion criteria were: [1] non-SCZ or FEP psychosis conditions or diagnoses including prodromal states (early-phase psychosis, clinical high-risk for psychosis, and sub-clinical psychosis), substance-induced psychosis, delusional disorder, and schizotypal personality disorder and [2] any exclusion of past year cannabis use (short–term cannabis washout before study evaluation was allowed).

### 2.3 Case Series Data

Psychiatric inpatients with auditory hallucinations were recruited for a feasibility study evaluating the response of mismatch negativity amplitude to psychosis treatment. For this report, we identified and compiled data for participants who reported cannabis use in the month prior to psychiatric hospital admission. Within one week after admission, trained researchers completed a structured interview to obtain demographics, symptoms, and past– month cannabis use. Psychotic symptoms were measured with the 6-item Positive and Negative Syndrome Scale for measurement of symptom severity (PANSS-6) (Østergaard et al., 2016). Cannabis use was assessed with the Cannabis Abuse Screening Test (CAST) and Cannabis Engagement Assessment (CEA) (Legleye, 2018; Schluter and Hodgins, 2022). The parent study was reviewed and approved by the New Hampshire Department of Health and Human Services Committee for the Protection of Human Subjects. All participants gave informed consent.

### 2.4 Cannabis Use Data

For comparability, where possible, we converted cannabis use amounts reported by case series participants and by participants in the reviewed studies into calculated daily joints using the mean estimated cannabis weight per joint of 0.32 grams (Ridgeway and Kilmer, 2016).

## 3. Results

### 3.1 Scoping Review Study Selection

The initial literature search resulted in 672 articles. Removing duplicates and articles published prior to 2016 (281) yielded 388 articles. Of these, 317 were excluded after screening and abstract review. Of the remaining 71 articles, a full–article review resulted in excluding 63 and retaining eight. We included an additional article published after the completion of the literature search that met inclusion criteria (Figure 1, Table 1).

**Table 1.** Methods and purposes of included studies analyzing cannabis use in schizophrenia-spectrum disorders.

| Author, Year | N total | Design | Location | Cannabis Assessment | Main Study Purpose |
| --- | --- | --- | --- | --- | --- |
| Studies comparing cannabis use to controls |  |  |  |  |  |
| Arranz 2018 | N = 207 | Cross sectional, observational trial<br>Enrolled likely after 2016 | Reus, Spain | Detailed team-designed clinical interview<br>Baseline assessment | Determine relationship between trauma, stress factors, and cannabis use exposure with psychosis in ROP and healthy controls |
| Brunette 2024 | N = 130 | Randomized, double-dummy, placebo-controlled trial<br>Enrolled 2014–2020 | New Hampshire and Vermont, US | Timeline Follow-Back method<br>Baseline assessment | Examine effects of oral or smoked THC vs placebo on brain reward, psychosis symptoms and cognition in SCZ–CUD and CUD only |
| Schnell 2019 | N = 108 | Controlled comparative study<br>Enrolled likely after 2016 | Köln, Germany | Structured interview with clinical verification<br>Baseline assessment | Investigate inhibition of return function as vulnerability marker in SCZ and healthy controls with or without cannabis use |
| Yeap 2023 | N = 55 | Cross sectional observational trial<br>Enrolled 2012–2021 | Toronto, Canada | Timeline Follow-Back method<br>Baseline assessment | Secondary analysis investigating effects of tobacco dependence and psychiatric diagnosis on cannabis withdrawal severity in SCZ–CUD and CUD only |
| Studies not comparing cannabis use to controls |  |  |  |  |  |
| Ghelani 2024 | N = 15 | Cross sectional qualitative interviews. Enrolled after 2018 | Ontario, Canada | Semi-structured interview<br>Baseline assessment | Investigate perspectives of young adult cannabis users with ROP in EPI towards CBD. |
| Gunasekera 2023 | N = 34 | Within-subject, crossover, double-blind, placebo-controlled trial<br>Enrolled 2016–2018 | London, UK | Research interview<br>Baseline assessment | Investigate CBD effects on motivational salience through fMRI measures of insular activation among patients with early psychosis |
| Madero 2020 | N = 106 | Observational cross-sectional<br>Enrolled Mar–Aug 2018 | Barcelona, Spain | Clinical interview<br>Baseline assessment | Investigate effects of cannabis use prior to inpatient admission on psychiatric severity among inpatients with SCZ and other diagnoses |
| Petros 2022 | N = 16 | Cross sectional qualitative, interviews<br>Enrollment likely after 2016 | Washington, US | Online screening<br>Baseline assessment | Identify factors for cannabis reduction or discontinuation in FEP |
| Tatar 2023 | N = 104 | Observational cross-sectional online survey of FEP patients<br>Enrollment: 2020–2022 | Quebec, Alberta, and Nova Scotia, Canada | Non-validated survey<br>Baseline assessment | Evaluate preferences of app-based interventions for treatment of CUD among FEP patients with CUD |

Key: THC = tetrahydrocannabinol, CBD = cannabidiol, SCZ = schizophrenia spectrum disorder, CUD = cannabis use disorder, SCZ–CUD = schizophrenia and cannabis use disorder, EPI = early psychosis intervention, FEP = first episode psychosis, ROP = recent-onset psychosis, fMRI = functional magnetic resonance imaging, MRI = magnetic resonance imaging

**Figure 1.**
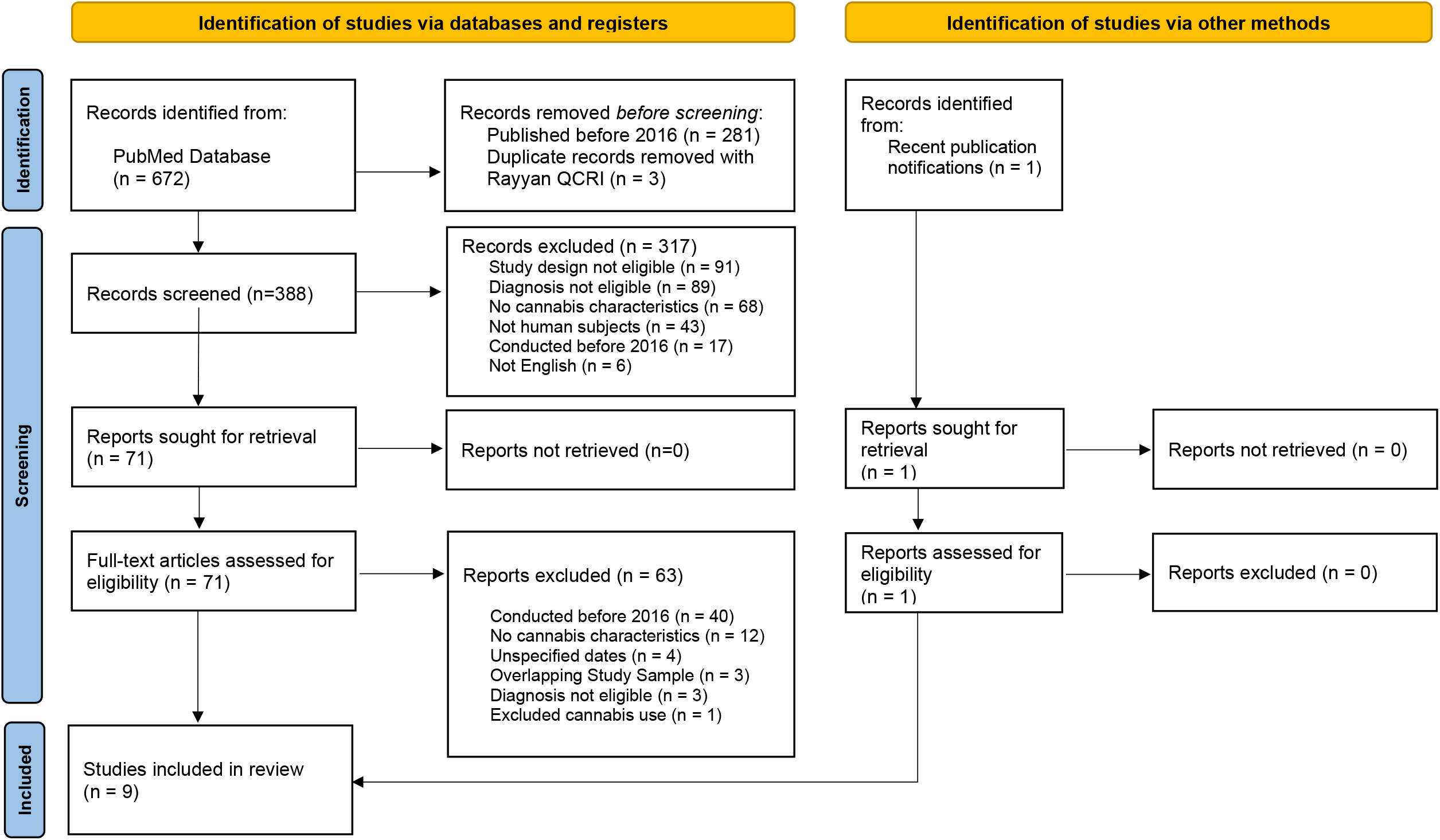
PRISMA diagram for review screening strategy. Of the 672 studies, nine were included in the scoping review.

### 3.2 Study Characteristics

Nine studies with various methodologies and purposes met inclusion/exclusion criteria (Table 1). None were designed to characterize cannabis quantity, frequency, or type of use over time. Study purposes ranged from qualitative interviews to assess cannabis use and perspectives on smartphone applications for treatment to functional imaging assessment of CBD effects on brain measures of motivational salience. Cannabis assessments were used to characterize participants to investigate a range of research questions. Among studies with control groups, four included people with cannabis use and without psychosis as a control condition (three studies with CUD, one with any use) (Arranz et al., 2018; Brunette et al., 2024; Schnell et al., 2019; Yeap et al., 2023). Of these four studies, three were controlled observational studies (Arranz et al., 2018; Schnell et al., 2019; Yeap et al., 2023) and one was a randomized, double-blinded, and placebo-controlled trial (Brunette et al., 2024). Of the five studies that did not include cannabis using comparison groups, four used observational designs (Ghelani, 2024; Madero et al., 2020; Petros et al., 2022; Tatar et al., 2023), two of which employed qualitative interview thematic analysis (Ghelani, 2024; Petros et al., 2022). The fifth study was a randomized trial among SCZ cannabis users (Gunasekera et al., 2023). The primary outcome measures were heterogeneous (Table 1). Cannabis use was assessed with unstructured clinical assessments, structured interviews, and the Timeline Follow Back (TLFB) method. Only one study utilized standardized interview technique - the TLFB method to obtain “typical joints” per day (Brunette et al., 2024). Another used “ad hoc questionnaire and chart review” but standardized the amount of cannabis using the Standardized Joint Unit (SJU) (Madero et al., 2020). None of these recent studies reported cannabis use characteristics over time (baseline and follow-up).

Four studies were based in Europe (Arranz et al., 2018; Gunasekera et al., 2023; Madero et al., 2020; Schnell et al., 2019), three in Canada (Ghelani, 2024; Tatar et al., 2023; Yeap et al., 2023), and two in the U.S. (Brunette et al., 2024; Petros et al., 2022). Only five studies required that participants used cannabis (Brunette et al., 2024; Ghelani, 2024; Petros et al., 2022; Schnell et al., 2019; Yeap et al., 2023).

The cumulative total of participants from all studies was 775. We based our results on the 432 participants diagnosed with SCZ, including FEP and ROP. 265 participants were engaged in early intervention or first–episode psychosis treatment. 106 participants were inpatients. Sample sizes ranged from 15 (Ghelani, 2024) to 207 (Arranz et al., 2018) across the included studies. Seven studies had samples with mean age under 30 years and two studies had samples with mean age around 35 (Madero et al., 2020; Yeap et al., 2023).

### 3.3 Cannabis Use Patterns Among People with SCZ in Clinical Research

#### 3.3.1 Frequency and Amount of Cannabis Use

Frequency of cannabis use was the most consistently reported pattern variable across studies and amount of use was the second most common. Among the three studies that did not require daily cannabis use for inclusion; two reported that 41.1%–66.7% of SCZ participants used weekly or more (Arranz et al., 2018; Gunasekera et al., 2023). Data from the three studies that reported amounts of use were converted into standard joints per day (Brunette et al., 2024; Schnell et al., 2019; Yeap et al., 2023) (Table 2). These studies showed that mean amounts of daily use among people with SCZ and CUD ranged widely with large standard deviations from 0.6 ± 0.6 (Brunette et al., 2024) to 3.4 ± 2.2 joints per day (Yeap et al., 2023). This wide variability in amount of use was also clearly seen in a Spanish study that reported participants with SCZ (without requiring CUD) used 0.9 ± 1.8 joints per day during the week prior to inpatient admission (Madero et al., 2020).

**Table 2.** Cannabis use characteristics in schizophrenia-spectrum disorders in studies recruiting since 2016.

| Author, Year | Group, Sample size | Cannabis Use Required | Cannabis Measures | Frequency or Amount of Use | Modality of Use | Use Legal<sup>a</sup> |
| --- | --- | --- | --- | --- | --- | --- |
| Studies comparing cannabis use to controls |  |  |  |  |  |  |
| Arranz 2018 | ROP *n* = 146<br>HC *n* = 61 | No | Prior 6-month frequency of use (% using >weekly to daily) | ROP vs. HC:<br>41.1% vs. 13.1% used ≥ weekly or more | NR | No |
| Brunette 2024 | SCZ–CUD *n* = 34<br>CUD only *n* = 51 | Yes, CUD | Days of any cannabis use and calculated mean joints/day in 35 days prior to enrollment<sup>b</sup> | SCZ–CUD vs. CUD only:<br>14.7 ± 10.1 vs. 26.4 ± 11.5 days/35 days<br>0.6 ± 0.6 vs. 1.4 ± 1.7 joints/day | Smoke | Yes |
| Schnell 2019 | SCZ–CUD *n* = 32<br>CUD *n* = 16 | Yes, ≥ once monthly | Mean joints used per month, calculated mean joints/day | SCZ–CUD vs. CUD only:<br>10.7 ± 15.3 vs. 13.7 ± 14.1 joints/month<br>0.4 ± 0.5 vs. 0.5 ± 0.5 joints/day | NR | No |
| Yeap 2023 | SCZ–CUD *n* = 35<br>CUD *n* = 20 | Yes, CUD | Mean grams/day use in last week, calculated mean joints/day<sup>c</sup> | SCZ–CUD vs. CUD only:<br>1.1 ± 0.7 vs. 1.4 ± 1.1 g/day<br>3.4 ± 2.2 vs. 4.3 ± 3.4 joints/day | NR | Partial |
| Studies not comparing cannabis use to controls |  |  |  |  |  |  |
| Ghelani 2024 | ROP *n* = 15 | Yes, ≥ daily | Current and lifetime frequency of use | THC = 54% used > 3 times daily<br>CBD = 33% reported lifetime use > 4–50 times | Flower 93%<br>Edible 80%<br>Concentrate 60%<br>Vape 40% | Yes |
| Guna-sekera 2022 | SCZ *n* = 15 | No | Past and current frequency of use | 66.7% used weekly to daily | NR | No |
| Madero 2020 | SCZ *n* = 35 | No | SJU and calculated mean joints/day in week prior to admission | 8.3 ± 16.4 Standard Joint Units<br>0.9 ± 1.8 joints/day | NR | No |
| Petros 2022 | SCZ *n* = 16 | Yes | Prior 1-month days of use and highest lifetime use period | 11.8 ± 12.5 days used in past month<br>0.9 ± 1.8 days used in highest use period | NR | Yes |
| Tatar 2023 | ROP *n* = 104 | Yes, CUD | Prior 3-month frequency of use | 43.3% used daily<br>30.8% used 3–6 days per week | NR | Yes |

Key: ROP = recent onset psychosis, HC = healthy control, SCZ = schizophrenia spectrum disorder, CUD = cannabis use disorder, NR = not reported, SJU = standard joint units
<sup>a</sup> Location of study included states with legal medical or recreational cannabis
<sup>b</sup> Calculated daily joints are based on the total 35-day collection period and not only on days of use
<sup>c</sup> Mean grams per day used were obtained from the paper's supplementary materials

**Table 3.** Case descriptions of people with psychosis and cannabis use.

|  |  |
| --- | --- |
| Case 1 | White, non-Hispanic male in his 50s with SCZ. PANSS-6 score was 22. He initiated cannabis between ages 5-10 and began to use regularly between ages 10-15. In the past month, he reported using cannabis leaf through pipes/one-hitters and typically using 2 joints each day for 14 of the past 30 days. |
| Case 2 | White, non-Hispanic female in her 20s with BPAD type 1 with psychosis. PANSS-6 score was 16. She initiated cannabis between ages 10-15 and began to use regularly between ages 15-20. In the past month, she reported using cannabis leaf once through joints/blunts, using 0.25 grams (0.78 calculated daily joints). In the past month, she also reported using cannabis concentrate budder through dabbing on 9 days, typically using one drop (1 mg of THC per 1 mL). |
| Case 3 | White, non-Hispanic male aged 15-20 years old with SCZ-F. PANSS-6 score was 15. He initiated cannabis between ages 10-15 and began to use regularly between ages 15-20. In the past month, he reported using on 14 days. He used cannabis leaf with 28% THC through joints/blunts and typically used 1.5 g (4.69 calculated daily joints) per day. In the past month, he also reported using an unspecified amount of cannabis concentrate shatter with a bong on 13 days. |
| Case 4 | Multiracial (black and white) non-Hispanic female aged 15-20 years old with SCZ-F. PANSS-6 score was 12. She initiated cannabis between ages 15-20 and began to use regularly after 1 year. In the past month, she reported using cannabis leaf on 1 day through joints/blunts and used 0.5 g (1.56 calculated daily joints) per day. In the past month, she also reported using an unspecified amount of cannabis candy on 1 day. |
| Case 5 | White, non-Hispanic male in his 30s with SCZ. PANSS-6 score was 15. He initiated cannabis between ages 15-20 and began to use regularly between ages 20-25. In the past month, he reported using cannabis leaf strain of Holland's Hope with 16% THC through an unspecified modality almost daily (25 of 30 days) and typically used 2 g (6.25 calculated daily joints) per day. |
| Case 6 | White, non-Hispanic male in his 30s with unspecified SCZ. PANSS-6 score was 15. He initiated cannabis between ages 20-25 and began to use regularly after 2 years. In the past month, he reported using cannabis leaf through pipes/one-hitters of an unspecified amount per day for 14 days. |

Key: SCZ = schizophrenia, SCZ-A = schizoaffective disorder, SCZ-F = schizophreniform disorder, HS = high school, PANSS-6 = 6-item Positive and Negative Syndrome Scale, BPAD = bipolar affective disorder, THC = tetrahydrocannabinol

#### 3.3.2 Cannabis Use in SCZ Compared to CUD without Psychosis

Among the three studies with comparison participants with CUD, no consistent pattern was observed in amounts of use between participants with SCZ and CUD and CUD without psychosis. We calculated mean joints from three studies (Germany, Canada, U.S.) – two showed no difference between groups (Schnell et al., 2019; Yeap et al., 2023) and the one U.S. study showed less frequent use (about half as many days in the last 35 days) among participants with SCZ and CUD compared to participants with CUD only (Brunette et al., 2024).

#### 3.3.3 Age of Cannabis Initiation

In one study, the age of cannabis initiation was significantly earlier in SCZ with CUD (15.81 ± 2.26 years) compared to participants with CUD without psychosis (18.04 ± 2.15 years) (Schnell et al., 2019). In another FEP sample (Canada), the mean age of cannabis initiation was also 15.27 ± 2.78 years (Tatar et al., 2023). One study reported no difference in the duration of cannabis use in SCZ with CUD (85.88 ± 46.6 months) and similarly aged participants with CUD without psychosis (98.64 ± 50.1 months) (Schnell et al., 2019).

#### 3.3.4 Cannabis Use Modality and Product Types

Smoking leaf cannabis was the most commonly reported modality and type of use (Brunette et al., 2024; Ghelani, 2024). Two studies described their samples as “smokers” but did not formally assess modalities of cannabis use (Arranz et al., 2018; Yeap et al., 2023). Only one small study comprehensively assessed participants’ lifetime THC and CBD product types and modalities among daily cannabis users with SCZ (Ghelani, 2024). The most used products were flower (93.3%), edibles (80%), and concentrates (60%) for THC and flower (40%) for CBD, with topicals and oils used less frequently for CBD. Modalities of use included smoking, vaping, ingestion, and topical application.

### 3.4 Case Series of Cannabis Use Patterns for Schizophrenia-Spectrum Disorders

Among the six psychiatric inpatients with clinically observed psychotic symptoms recruited in 2023–2024 (Table S3), the mean age was 32.00 ± 14.42. Most were male (66.7%), white (83.3%), and diagnosed with SCZ (83.3%). The mean PANSS-6 score was 15.83 ± 3.31 and the average cannabis initiation age was 14.17 ± 4.88 years.

Participants reported using leaf, concentrate (shatter, budder), and edible (gummy, mint) products (Table S4). All patients used cannabis leaf products (mean daily joints 3.06 ± 2.31, mean use days per week 2.68 ± 2.14) and 66.7% reported heavy use (≥2 joints daily). Half of the participants (all heavy users) also used concentrates (33.3%) and edibles (16.7%). Modalities of cannabis use included pipes and joints for leaf products and bongs and dabs for concentrates. All participants used cannabis mostly alone and half obtained cannabis from a dealer. Most (83.3%) reported they had tried to reduce use, but none had sought formal or informal support for reduction.

## 4. Discussion

Our scoping review of recent research on specific cannabis use patterns in clinical populations with SCZ yielded only nine studies, all with relatively small study samples. None of the studies were specifically designed to characterize cannabis use in this population. Instead, all studies described cannabis use in the context of testing an array of hypotheses. The type and quality of measures used to characterize cannabis use varied widely such that studies were difficult to compare. No studies reported on frequency, quantity, or type of cannabis use over time in SCZ. Based on this review, there is insufficient recent research to confidently characterize cannabis use patterns in clinical populations of persons with SCZ, particularly in the era of cannabis legalization in the U.S., thus further research is needed.

Regarding measurement of cannabis use, only one study used a standand interview technique (Brunette et al., 2024). In order to standardize measurement of cannabis in research participants, prior consensus from expert cannabis researchers had proposed the International Cannabis Toolkit (iCannToolkit), which focuses on three heirarchal layers – universal measures, detailed self-report, and biological measures (Lorenzetti et al., 2022). Recommendations included basic universal measures of cannabis frequency, Timeline Follow-Back (TLFB) methodology, and THC measurements in urine, saliva, and plasma, measures which vary in cost and ease of implementation. Given impaired recall accuracy from psychotic and cognitive symptoms, quantitative biological measures (e.g. urine or serum THC metrics) may confirm recent self-reported cannabis use in persons with psychotic disorders, although the storage and delayed release of THC can create challenges to interpreting such measures (Chesney et al., 2024). Current research is working to establish better strategies to measure cannabis consumption behavior and estimate the quantity and type of consumed cannabis (Borodovsky et al., 2023; Budney et al., 2022; Walsh et al., 2023).

The nine recent studies reviewed here demonstrated a range of self-reported cannabis use in participants with SCZ. Some participants used cannabis infrequently, but a substantial proportion reported using heavily and used high-potency products. In the case series of inpatients with psychosis and recent cannabis use, most were frequent, heavy users and half used multiple products. Similar to research enrolling FEP subjects prior to 2016, these findings suggest that a subset of people with SCZ use frequently and heavily and highlight that some use high–potency cannabis products (Quattrone et al., 2021). These findings in clinical samples align with the results of the previously mentioned recent online survey of a non-clinical sample (Rup et al., 2021).

Overall, our review found little recent research that characterizes cannabis use in SCZ: other recent U.S. research related to legalization has focused on the harms of cannabis use regarding psychosis. One study used national claims data and found no significant association between state cannabis legalization and psychosis-related health outcomes (Elser et al., 2023). A Colorado study reported higher rates of emergency department use for psychosis but not for SCZ in counties with increases in recreational cannabis dispensaries (Wang et al., 2022). Additionally, the recent survey noted above also found that perceived harms and help–seeking for harms related to cannabis use were more likely to be reported by those with self–reported psychosis compared to those without mental health conditions (AOR = 1.78; 1.11–2.87) (Rup et al., 2022). Additional research has examined impacts of legalization in Canada. A recent Canadian population cohort study showed a tripled proportion of incident SCZ cases associated with CUD from the pre–legalization (2006–2015; PARF for CUD = 3.7%; 2.7–4.7%) to the post–legalization (2018–2022; PARF for CUD = 10.3%; 8.9–11.7%) periods (Myran et al., 2025). Additionally, development of SCZ was more likely in persons with CUD (8.9%) compared to those without CUD (0.6%). The incidence of unspecified psychosis nearly doubled between pre–legalization and post–legalization periods (30 to 55.1 per 100,000 individuals, respectively). These early impacts of cannabis legalization highlight the need for further understanding of cannabis use patterns in highly vulnerable populations like SCZ and long–term impacts of legalization on psychosis outcomes.

Similar to recent research, most past research (e.g. prior to the era of legalization) was also not primarily designed to describe cannabis use quantity, type, and modality in SCZ populations. One study estimated cannabis consumption using self–reported joint equivalents of Δ9-tetrahydrocannabinol (Δ9-THC) form of cannabis (0.5 grams estimated to be equivalent to 2–4 joints of Δ9-THC) and weekly cost of cannabis consumption but did not report cannabis use specific amounts (Jockers-Scherübl et al., 2007). Cannabis use frequency and quantity was reported in one small study sample in which participants with SCZ and CUD reported 4.5 ± 2.1 use days and 12.3 ± 12.0 joints used per week (Brunette et al., 2011), but data on cannabis type was not reported. Additionally, thresholds for heavy cannabis use were lower compared to current studies. For example, in one study sample enrolled between 2002–2006, “high use” was defined as 24 or more incidents of use in the last 24 months (Ringen et al., 2008).

Several limitations to our review and case series should be noted. This study utilized a single database (PubMed only) and a small number of cases are reported in the case series. It is possible that other studies meeting our inclusion criteria are only available from other literature databases. Yet this review clearly demonstrates an interesting gap in recent evidence regarding patterns of cannabis use in SCZ, a substance with likely harms to a vulnerable clinical population. This gap is particularly compelling as many states and the U.S. as a country consider expanding legalization of cannabis.

## 5. Conclusion

As cannabis legalization proceeds in the U.S. and internationally, it is crucial to understand how vulnerable populations may be impacted. Little is known about the characteristics of cannabis use in persons with SCZ. Among the recent nine studies we identified, samples were relatively small, most lacked standardized or recommended measures, and studies were not primarily designed to determine cannabis use characteristics in SCZ. These studies reported a range of cannabis use amounts and frequencies in persons with FEP and SCZ with some showing substantial proportions of participants who used daily and/or heavily. Two–thirds of individuals in the case series were heavy users and half used concentrates or edibles in addition to leaf products. As cannabis legalization expands, further research should characterize cannabis use in SCZ and examine the extent of risky high-potency product use in people with primary psychotic disorders.

## Data Availability

All data produced in the present work are contained in the manuscript

## 6. Acknowledgements

The case series was part of a feasibility study funded by the Matthew Friedman – Gary Tucker Junior Researcher Award provided by the Department of Psychiatry, Dartmouth-Hitchcock Medical Center. We would like to acknowledge Jenna Bourassa for her crucial role in providing study coordination and project feedback.

## 7. Author Contributions

**Jeff W. Jin:** Methodology, Investigation, Formal Analysis, Writing – Original Draft, Review & Editing, Visualization. **Nolan J. Neu:** Investigation, Writing – Original Draft, Review & Editing. **Isaac B. Satz:** Writing – Original Draft. **Janelle Annor:** Investigation. **Mohamed ElSayed:** Data Curation (Case Series), Writing – Review & Editing. **Mary F. Brunette:** Conceptualization, Methodology, Investigation, Writing – Original Draft, Review & Editing, Visualization, Supervision.

