## Supplementary Materials for "Cannabis Use Patterns in First Episode Psychosis and Schizophrenia: A Scoping Review and Case Series"

**S1. PubMed Literature Search Design**

- (“cannabis”[mesh] OR “cannabinoids”[mesh] OR “tetrahydrocannabinol”[tiab] OR “THC”[tiab] OR “CBD”[tiab])

  **AND**

- (“schizophrenia spectrum and other psychotic disorders”[mesh] OR “deficit syndrome”[tiab] OR “deficit schizophrenia”[tiab] OR "severe mental illness"[tiab] OR "SMI"[tiab])

  **AND**

- (“product”[tiab] OR “amount”[tiab] OR “potency”[tiab] OR “motiv*”[tiab] OR “duration”[tiab] OR “epidem*”[tiab] OR “public health”[tiab] OR “pattern“[tiab] OR “prevalence”[tiab] OR “incidence”[tiab] OR “legalization”[tiab] OR “outcome”[tiab] OR “course”[tiab] OR “function”[tiab] OR “deficit”[tiab] OR “hospital admission”[tiab] OR “exacerba*”[tiab] OR “improv*”[tiab] OR “increas*”[tiab] OR “decreas*”[tiab] OR “chang*”[tiab])

**S2. Scoping Review Inclusion and Exclusion Criteria**

Inclusion Criteria:

[1] Publication in English language

[2] Inclusion of individuals with Diagnostic and Statistical Manual of Mental Disorders, Fourth Edition (DSM-IV), or revised diagnosis of SCZ (schizophrenia, schizoaffective disorder, schizophreniform disorder) including chronic SCZ, first episode psychosis [FEP], or recent-onset psychosis [ROP])

[3] Recruitment of subjects resulting in sample with at least half of data collected after 2016

[4] Systematic assessment of cannabis use, frequency, or other characteristics

Exclusion Criteria:

[1] non-SCZ or FEP psychosis conditions or diagnoses including prodromal states (early-phase psychosis, clinical high-risk for psychosis, and sub-clinical psychosis), substance-induced psychosis, delusional disorder, and schizotypal personality disorder

[2] any exclusion of past year cannabis use (short–term cannabis washout before study evaluation was allowed)

[3] Lack of SCZ diagnosis at study initiation

[4] Report of only CUD diagnosis without further cannabis characteristics

**Table S3.** Demographics of case series participants experiencing psychosis and reporting cannabis use in the past month

| | **PSY CN 1M** (*N* = 6) |
|---|---|
| Schizophrenia diagnosis (%) | 5 (83.3) |
| Age, mean (SD) | 32.00 (14.42) |
| Birth Sex, male (%) | 4 (66.7) |
| Race, white (%) | 5 (83.3) |
| Ethnicity, not Hispanic or Latino (%) | 5 (83.3) |
| Total Education (%) | |
| Completed ≤ high school | 5 (83.3) |
| Completed > high school | 1 (16.7) |
| Major Income Source (%) | |
| Self | 5 (83.3) |
| Other (SSD) | 1 (16.7) |
| Total PANSS-6 score (SD) | 15.83 (3.31) |
| Estimated daily tobacco use in past month, cigarettes (%) | |
| Non-smoker | 2 (33.3) |
| Light to Moderate (0 – 20) | 4 (66.7) |

*PSY CN 1M = psychosis with cannabis use in the past month

**Table S4.** Cannabis characteristics of case series participants experiencing psychosis and reporting cannabis use in the past month

| | **PSY CN 1M** ($N$ = 6) |
|---|---|
| **General Cannabis Characteristics** | |
| Age of first cannabis use, mean (SD) | 14.17 (4.88) |
| Age of initial occasional cannabis use[A], mean (SD) | 13.33 (11.38) |
| Total years of occasional cannabis use, mean (SD) | 5.17 (9.41) |
| Age of initial regular cannabis use[B], mean (SD) | 18.67 (4.46) |
| Total years of regular cannabis use, mean (SD) | 11.67 (14.42) |
| Separate days of any cannabis product use in the past month, mean (SD) | 10.33 (9.40) |
| Social setting of cannabis use (%) | |
| Always or almost always with alone | 6 (100) |
| Method of cannabis obtainment (%) | |
| Dealer | 3 (50) |
| Home grown | 1 (16.67) |
| Government-run cannabis store (online, in-person) | 2 (33.33) |
| Concurrent substance use (%) | |
| Alcohol | 1 (16.67) |
| Tobacco | 1 (16.67) |
| Attempts to reduce cannabis use (%) | |
| Yes | 5 (83.33) |
| No | 1 (16.67) |
| Sought formal or informal support[C] to reduce cannabis use (%) | |
| No | 6 (100) |
| **Leaf Cannabis Products** | |
| Individuals reporting use in past month (%) | 6 (100) |
| Modality of use (%) | |
| Pipe/One-hitter | 2 (33.33) |
| Joint/Blunt | 3 (50) |
| Other (unspecified) | 1 (16.67) |
| Amount of daily use in the past month (%) | |
| Less than 2 joints daily | 2 (33.33) |
| Greater than or equal to 2 joints daily | 4 (66.67) |
| Mean joints used per day, mean (SD) | 3.06 (2.31) |
| Mean days of use per week, mean (SD) | 2.68 (2.14) |
| Use of any other non-leaf products | 3 (50) |
| **Concentrated Cannabis Products** | |
| Individuals reporting use in past month (%) | 2 (33.33) |
| Modality of use (%) | |
| Bong | 1 (50) |
| Dabbing | 1 (50) |
| Product Type (%) | |
| Shatter | 1 (50) |
| Other: Budder | 1 (50) |
| **Edible Cannabis Products** | |
| Individuals reporting use in past month (%) | 1 (16.67) |
| Product type (%) | |
| Candy (gummies, mints) | 1 (100) |

*PSY CN 1M = psychosis with cannabis use in the past month

[A]One to two times monthly
[B]Greater than weekly
[C]From a physician, health provider, counselor, or informal support
